# The age-specific concentrations and seroprevalence of antibodies against *Salmonella* Enteritidis (O:9) and *Salmonella* Typhimurium (O:4,5) across three sites in Kenya

**DOI:** 10.1101/2025.04.14.25325777

**Authors:** Esther M Muthumbi, Sean C Elias, Alfred Mwanzu, Agnes Mutiso, Perpetual Wanjiku, Cecilia Mbae, Godfrey Bigogo, Jennifer R. Verani, Stefan Flasche, Samuel Kariuki, Calman A. MacLennan, J Anthony G Scott

## Abstract

**Background:** Seroprevalence studies can indicate the age-distribution of first infection with non-typhoidal *Salmonella* (NTS) and estimate the rate of infection by age. This can help target interventions, particularly vaccination.

**Method:** We collected 1254 paired samples of serum and stool from healthy children and adults (aged 0-82 years) in Kilifi, Nairobi, and Siaya counties in Kenya, areas of low, medium and high incidence of invasive NTS disease (iNTS), respectively. We quantified the age and site-specific geometric mean concentrations (GMCs) of IgG and IgA antibodies against *S*. Enteritidis O-antigen (O:9, serogroup D) and *S*. Typhimurium O-antigen (O:4,5, serogroup B) using an in-house standardised ELISA. A previously calibrated reference serum was used as an internal control. The threshold for seropositivity was determined using mixture modelling. Stool samples were cultured for NTS; isolates were serotyped using the Kauffman-White scheme.

**Results:** Maternally derived O:9 IgG and O:4,5 IgG antibodies were detectable in 100% of neonates and the GMC decreased by 40% (95%CI [25%-52%]) per month in the first six months of life. GMCs of IgA were low in neonates. After age 6 months, the O:9 and O:4,5 IgG and IgA GMCs increased sharply with age across all sites, reaching a plateau in early adulthood. The rate of increase in IgG GMCs by age was highest for O:9 in Nairobi, and for O:4,5 in Kilifi. Mixture modelling defined a threshold of 14.1 AU for O:9 IgG (87% sensitivity and 87% specificity) and 28.2 AU for O:4,5 IgG (89% sensitivity and 66% specificity). Seroprevalence also increased by age. The GMCs of O:4,5 IgG were 2 times higher for carriers of serogroup B *Salmonella* than non-carriers (GMC Ratio 2.4 95%CI [1.1-5.4]).

**Discussion:** Maternal antibodies to non-typhoidal Salmonella decay rapidly from 0-5months after which incident infection with both serogroups occurs. Therefore, control efforts should be implemented in early infancy before primary infection.

**Author Summary:** We have used serology to describe the epidemiology of NTS infection and rates of infection across three sites in Kenya with varying incidence of iNTS disease. Using a population-based, cross-sectional study, we have demonstrated that protective maternal antibodies are present at birth but decay within first 6 months of life after which antibody concentrations rise quickly with age implying a high rate of primary infections; therefore, infection control strategies need to be implemented in early infancy, and where vaccination is considered, the optimal age window for vaccination should consider the presence of maternal antibodies. In addition, we have highlighted areas that would benefit from further scientific research, including assay standardization and the thresholds for correlates of protection against infection and against invasive disease.

## Introduction

Invasive non-typhoidal Salmonella disease is a food-borne illness that causes significant morbidity and mortality globally, estimated as 535,000 cases and 77,500 deaths in 2017(1). The burden is highest in sub-Saharan Africa, in children <5 years old, and >80% is caused by two serovars: *S*. Typhimurium and *S*. Enteritidis (1, 2). Ingestion of non-typhoidal *Salmonella* (NTS) bacteria can lead to transient mucosal infection and clearance, asymptomatic colonisation, enterocolitis or invasive disease(3, 4). Hospital-based surveillance captures symptomatic cases, such as those with diarrhoea and those with bacteraemia, while community-based, cross-sectional studies among healthy individuals, estimate the prevalence of asymptomatic carriage(5, 6). Culture-based estimates of NTS burden are limited by the low sensitivity of culture for isolation of NTS. Additionally, hospital-based estimates are limited by access to diagnostic facilities, leading to underestimation of the burden of NTS. Quantification of IgG and IgA antibodies reflects the cumulative incidence of infection in the community(7, 8). Analysis of age-related serological profiles in respiratory syncytial virus (RSV), for example, can estimate the rate of acquisition of these infections by age and guide the optimal age group to target for interventions, particularly vaccination (9). For vaccines that might be designed to interrupt human-to-human transmission (10, 11), understanding the relationship between the antibodies and gut clearance is also critical.

There are currently no licensed vaccines against NTS. At present, there are several vaccine candidates against NTS in different stages of the vaccine development pipeline(9,10). Most target the serotypes responsible for the majority of iNTS cases, i.e. *S*. Typhimurium and *S*. Enteritidis, in bivalent formulations, or in multivalent formulations including typhoidal serotypes. These vaccines aim to prevent disease in infants, in whom disease incidence is highest (12). However, maternal antibodies may reduce the efficacy of infant vaccination(13). Assessment of the decay of maternally derived IgG antibodies in infants against the rate of acquisition of IgA antibodies in infants can identify the optimal window for vaccination with a future NTS vaccine(14).

There are no standardized assays for the quantification of anti-NTS antibodies (15) and no commonly agreed threshold for determining seropositivity, based on infection or on protection. Consequently, there are two analytic alternatives: 1) to analyse and characterise the data in their continuous form (16, 17) or; 2) to use modelling approaches that reconstitute unobserved populations (comprising uninfected and infected populations) given a set of reasonable assumptions(18, 19). This technique assumes that a given dataset contains unobserved groups within it and that these groups have distinct distributions and characteristics which can be inferred from the frequency distribution of antibody concentrations in the whole population. The technique has been used previously in infectious disease epidemiology to dichotomize populations into seronegative and seropositive groups with respect to malaria, dengue and SARS-COV2 antibodies(20-22), but has not been attempted for NTS.

The aim of this study was to estimate the age-specific geometric mean concentrations and seroprevalence of anti-NTS antibodies across three sites in Kenya with markedly different incidence of iNTS disease.

## Methods

A randomly selected, age-stratified sample of healthy children and adults from Kilifi, Nairobi and Siaya counties in Kenya were recruited into a community-based cross-sectional study of the prevalence of faecal carriage of NTS (n=1,497) conducted between October-November 2016 in Kilifi and May – September 2017 in Nairobi and Siaya (5). This involved collection of stool samples for isolation of NTS, anthropometry measures, and a questionnaire on WASH practices. Participants were also asked to provide a venous blood sample for testing of anti-NTS antibodies, spot testing for malaria parasites and haemoglobin concentrations. The samples were transported to the laboratory in cool-boxes at 4°, where the serum was separated, aliquoted and stored at −80°C.

### Laboratory methods

We used an indirect ELISA for detection of anti-O:4,5 antibodies representing *S*. Typhimurium and anti-O:9 antibodies representing *S*. Enteritidis (23). Purified O-antigens (CVD1925 and CVD1943) were used for plate coating (24). Nunc Maxisorb 96-well plates were coated with 5ug/ml O-antigen in coating buffer (5.3g Na_2_CO_3_ + 4.2g NaHCO_3_ + 1L dH_2_0, pH 9.6) and incubated overnight at +4⁰C. The plates were washed 5 times with phosphate-buffered saline (PBS) containing 0.05% Tween 20 (PBS-Tween) then blocked with Casein (Thermo Scientific, 37528) for 1 hour, and washed again 5 times with PBS-Tween and x1 with PBS. Test sera were initially diluted at 1:50 and plated in triplicates. Where higher dilutions were required, 1:500 or 1:5000 dilutions of the sera were used.

An in-house human control serum containing antibodies to the two antigens was used as reference sera for the standard curve (23, 25). These were plated in duplicate following a series of eleven 2-fold dilutions starting at 1:10 (or 1:20 for the O:4,5 IgG assay). Controls were included on each plate as follows: a negative control, consisting of casein dilution buffer added to 2 ‘blank’ wells; a positive control consisting of pooled serum from NTS-positive Malawian adults diluted in casein in 2 wells; an internal control, consisting of the reference sera diluted at 1:160 for IgA assays and 1:320 for IgG assays plated in 4 wells. The plates containing the test and reference sera were incubated for 2 hours at room temperature and subsequently washed with PBS-Tween x 5 and PBS x1. A corresponding secondary antibody, either the Anti-Human IgA (α-chain specific) peroxidase antibody produced in goat (Sigma, A0295) or the Anti-Human IgG (γ-chain specific) peroxidase antibody produced in goat (Sigma, A6029), was added to the plates which were then incubated at room temperature for 1 hour followed by a washing step with PBS-Tween x 5. The plates were developed using TMB Substrate (AbCam, ab171522) for the colour reaction, which was controlled by addition of the Stop solution (450nm TMB Stop Solution, AbCam ab171529). The development time was calibrated to generate an OD of 1 in the internal control. Once the Stop solution was added, the plates were read within 30 mins using ELISA reader operating Gen5 Software (version 2.0). Bound antibodies were quantified by measuring the absorbance at 450 nm and interpolating the values relative to the reference sera based on a 4-parameter logistic regression curve as ELISA Antibody Units (AU). One AU is equal to the reciprocal of the dilution of the standard serum giving an OD =1. Repeat testing of the samples was done if the coefficient of variation between the triplicates was >20%. Full plate repeats were done if the fit of the standard curve was unsatisfactory (R^2^<0.994), if the background reaction was high (negative control >0.15 OD) or if the Internal and Positive controls were not within the defined range. Additional dilutions of the samples were recommended if the OD values were higher than the top end of the reference curve, i.e. where the OD of the sample exceeds the OD of the 1^st^ dilution point of the standard curve. Those below the bottom end of the reference curve were considered negative. Stool samples were cultured on selective media after overnight incubation in Selenite F broth. *Salmonella* enterica species were confirmed using API 20E and thereafter, serotypes were determined using Kauffman-White scheme.

### Statistical Analysis

The antibody units were log-transformed (base 10) before further manipulation. Zero values were arbitrarily allocated a value that was half of the lower limit of quantitation (LLOQ) for the specific assay. Geometric mean concentrations (GMC) were estimated by age-strata used in sampling (0-11 months, 12-59 months, 5-14 years, 15-54 years and 55+ years), by site (Kilifi, Nairobi, Siaya) and by faecal NTS carriage status. Comparisons between groups were performed using ANOVA with Tukey’s pairwise comparison tests. We used the Reverse Cumulative Distribution (RCD) curves to visualize the distribution of antibody concentrations by age-group.

A piecewise regression model, with a single inflection point at age p, was used to assess the rate of change of antibody levels with age (x). It fit two linear regressions to data, one where x<p and the second one where x>=p. In Stata, this was implemented using the *nl* command. It estimates the inflection/breakpoint age p, the gradient in the age band where x<p, and the gradient in the age band where x>=p. To account for maternal antibodies, we excluded the period <6months (m>x>p).

Linear regression on log antibody concentrations was used to assess the mean rate of decay, d, of maternal antibodies, using the formula(26):

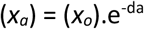

where (*x*_*a*_) refers to the mean antibody concentration at age a while (*x*_*o*_) refers to the mean antibody concentration at birth. These models were fit separately for each site.

A 2–component Gaussian mixture model was constructed to separate the data into two latent classes, assumed to be a seronegative class and a seropositive class. This approach uses the data to reconstruct the subclasses assuming that their distributions are normal and different from each other, deriving a mean and standard deviation (SD) for each sub-class. This was implemented using the *fmm* package in Stata(27). With the resulting distributions, we plotted a Receiver Operating Curve (ROC) which we used to estimate and compare the sensitivity and specificity of different cut-offs, including the Youden Index(28), assays’ LLOQ and the mean of the first class (assumed seronegative) +2SD. (19). Using the Youden Index as threshold, we assessed differences in seroprevalence by age and by site. All statistical analyses were conducted in Stata V15 (StataCorp, College Station).

## Results

Among 1497 participants from the parent survey, 1254 (84%) had a serum sample collected; of these, 319 (25%) participants came from Kilifi, 451 (36%) from Nairobi and 484 (39%) from Siaya. Due to limited volumes of sera, we assayed 1252 for O:9 IgG, 1216 for O:4,5 IgG and 1194 for O:9 and O:4,5 IgA (Supplementary Table S1). One sample had incomplete metadata and was excluded. 1148 samples had results for all 4 antigen-antibody combinations. Of the 1148 participants with complete data, the median age was 9.7 years (Interquartile range, 2.8-34 years) and 518 (45%) were male (Supplementary Table S2).

Both O:9 IgG and O:4,5 IgG were present among neonates and decreased to low levels by 4-5 months of life, before rising again (Figure 1). The proportion of neonates with detectable antibodies i.e. antibody units above the assays LLOQ, decreased from 100% to 71%, for O:9 IgG and from 100% to 74%, for O:4,5IgG between the first and 6^th^ month of life, respectively. The O:9 and O:4,5 IgG GMC among neonates was 20AU (95% CI [4-96]) and 23AU (95% CI [4-149]), respectively. The pooled rate of decay of both O:9 and O:4,5 IgG was 40% per month for the first 6 months of life. For IgA, the proportion of neonates with detectable O:9 and O:4,5 IgA was 67% and 33%, respectively, which increased to 94% and 78% at 5 months of life.

**Figure 1.**
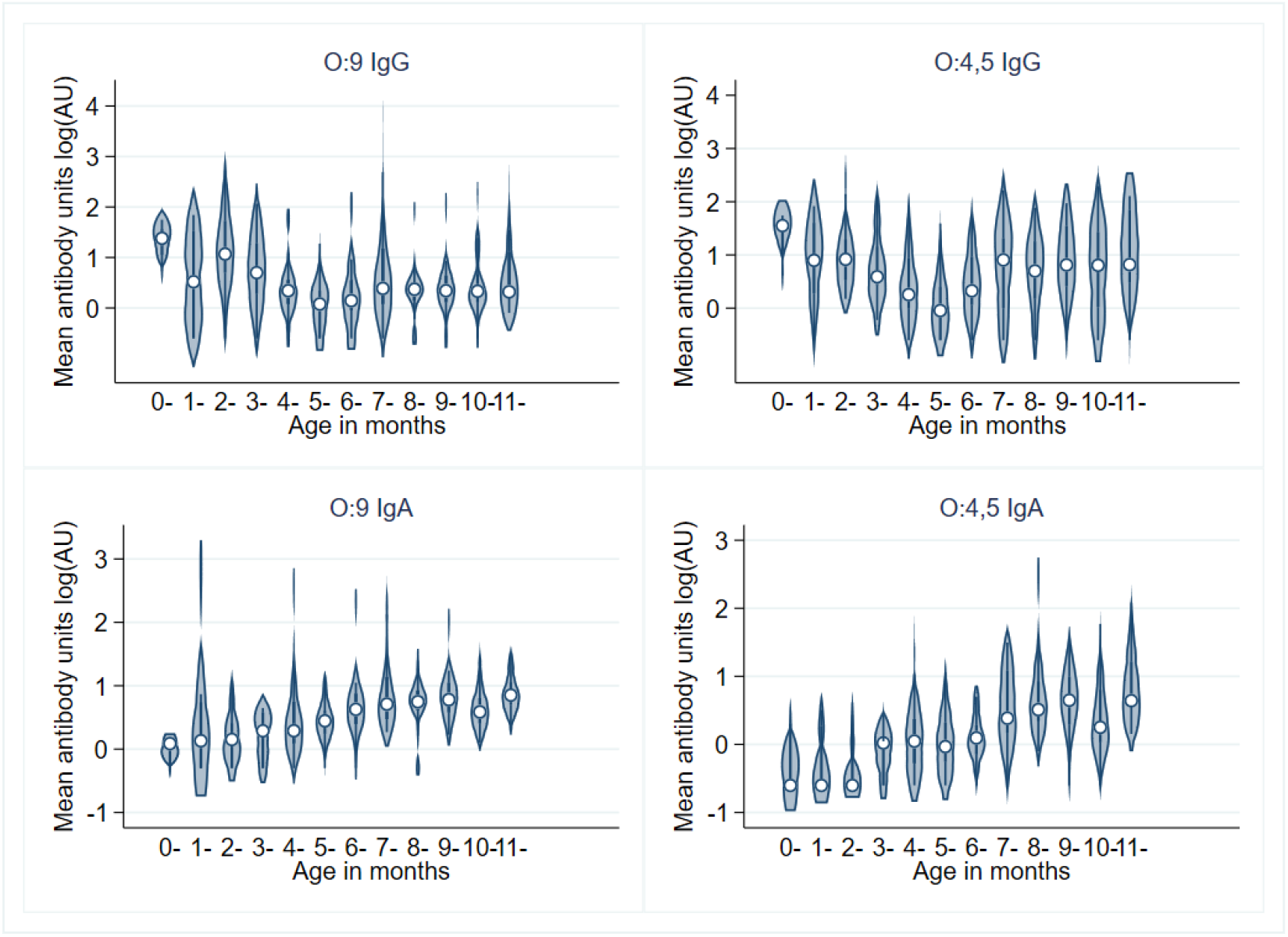
Decay of maternal antibodies (IgG) and early acquisition of antibodies (IgA) among infants.

After the first 6 months, The GMC for O:9 and O:4,5 IgG increased with age across all sites, shown by the shift to the right of the RCD curves with increasing age (Figure 2, Table 1).

**Table 1:** Geometric mean concentrations (95% CI) of antibody concentrations by age in Kenya.

|  |  | Kilifi |  | Nairobi |  | Siaya |  | All sites |  |
| --- | --- | --- | --- | --- | --- | --- | --- | --- | --- |
|  |  | GMC | 95% CI | GMC | 95% CI | GMC | 95% CI | GMC | 95% CI |
| O:9 IgG | 0-11 m | 6 | 4-11 | 2 | 2-4 | 3 | 2-4 | 3 | 3-4 |
|  | 12-59 m | 17 | 11-28 | 16 | 11-23 | 7 | 5-10 | 12 | 9-15 |
|  | 5-14 y | 25 | 16-38 | 28 | 21-37 | 23 | 18-32 | 26 | 21-31 |
|  | 15-54 y | 49 | 32-72 | 41 | 31-54 | 76 | 57-101 | 52 | 44-63 |
|  | 55+ y | 51 | 35-72 | 44 | 24-80 | 76 | 60-97 | 61 | 50-74 |
| O:4,5 IgG | 0-11 m | 10 | 6-16 | 3 | 2-4 | 5 | 3-7 | 5 | 4-6 |
|  | 12-59 m | 66 | 46-93 | 12 | 8-18 | 23 | 16-35 | 23 | 18-30 |
|  | 5-14 y | 76 | 56-101 | 44 | 35-56 | 60 | 45-80 | 54 | 46-64 |
|  | 15-54 y | 108 | 81-143 | 80 | 63-103 | 108 | 85-138 | 95 | 82-111 |
|  | 55+ y | 73 | 56-63 | 75 | 46-123 | 99 | 83-118 | 85 | 74-99 |
| O:9 IgA | 0-11 m | 3 | 2-4 | 3 | 2-4 | 7 | 5-9 | 4 | 3-5 |
|  | 12-59 m | 10 | 8-13 | 13 | 11-16 | 12 | 10-15 | 12 | 10-13 |
|  | 5-14 y | 17 | 14-22 | 23 | 20-28 | 26 | 20-33 | 23 | 20-26 |
|  | 15-54 y | 48 | 36-62 | 40 | 33-48 | 60 | 47-77 | 48 | 42-55 |
|  | 55+ y | 46 | 36-60 | 56 | 41-76 | 58 | 46-74 | 53 | 46-62 |
| O:4,5 IgA | 0-11 m | 2 | 1-3 | 2 | 1-3 | 2 | 2-3 | 2 | 1-2 |
|  | 12-59 m | 15 | 11-22 | 10 | 7-12 | 11 | 8-15 | 11 | 10-14 |
|  | 5-14 y | 19 | 15-26 | 20 | 16-24 | 27 | 21-35 | 22 | 19-25 |
|  | 15-54 y | 43 | 34-54 | 52 | 41-64 | 73 | 57-94 | 55 | 48-64 |
|  | 55+ y | 42 | 32-55 | 73 | 46-116 | 84 | 67-105 | 65 | 55-76 |

**Figure 2.**
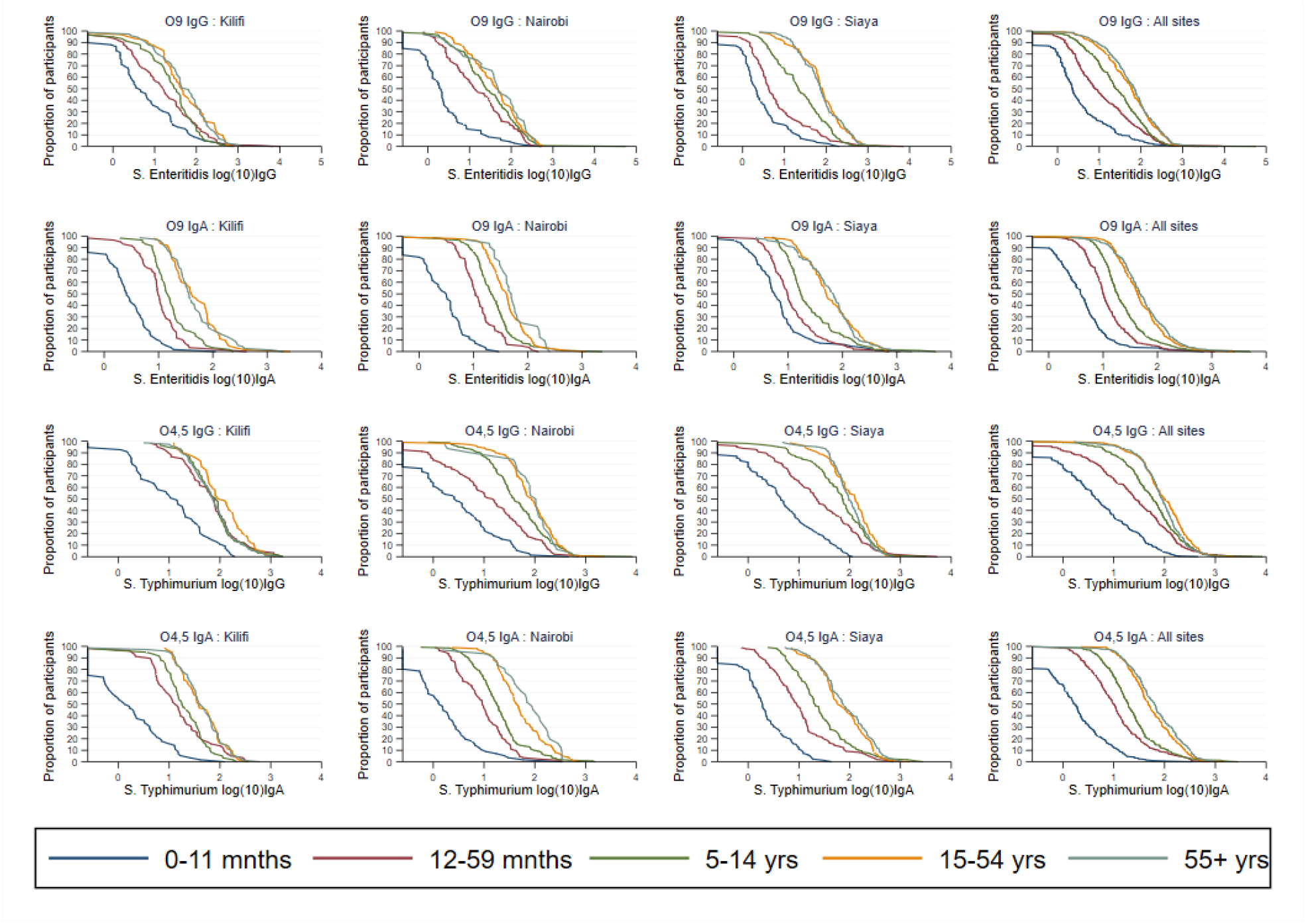
Reverse cumulative distribution curves of the concentrations of O:9 and O:4,5 IgG and IgA by age and by site.

A scatterplot (with LOWESS curve) of the concentration by age showed an increase during childhood, which peaked at a certain age (p) for each antibody specificity/isotype and then remained flat throughout adulthood (Supplementary Figure 1-2). To model this function, we fit a piecewise linear regression model with one inflection point, p, and estimated the rate of increase in antibody concentrations. Mean O:9 IgG increased at a faster rate in Nairobi than in Kilifi and Siaya, to reach its maximum concentration by 2.1 years while mean O:4,5 IgG increased fastest in Kilifi (Table 2). In both Kilifi and Siaya, the rate of increase in O:4,5 IgG was faster than O:9 IgG.

**Table 2:**
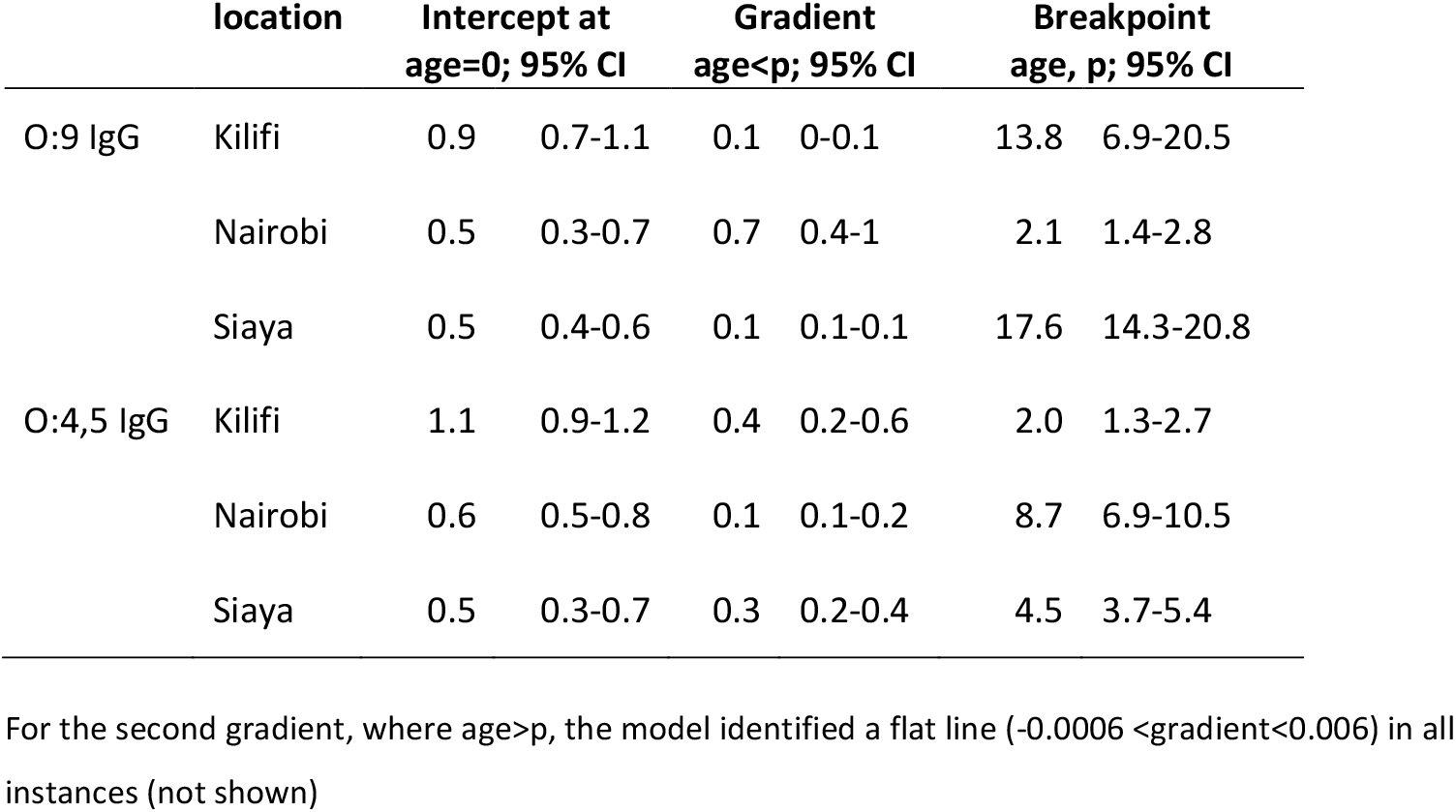
Piecewise regression analysis of the rate of increase in antibody concentrations by age in years at each site.

Among 1253 matched stool samples, 35 (3%) were positive for NTS culture. There were 6 serogroup D *Salmonella* isolates (4 were *S*. Enteritidis) and 10 serogroup B *Salmonella* isolates (1 was *S*. Typhimurium). Nineteen of the isolates could not be fully typed by the anti-sera available; however, they were neither serogroup B nor serogroup D. Of 35 NTS isolates, 28 (80%) were from Kilifi and 6 (17%) from Siaya. In Nairobi, only one stool sample was positive for NTS, and this was of an undefined serogroup. Among children aged < 5 years, 3 carried serogroup D while none carried serogroup B *Salmonella*.

GMCs of both IgG and IgA were higher for homologous carriers than non-homologous-carriers across the ages though the differences were not significant, except for participants aged ≥5 years where those with serogroup B carriage had 2 times higher O:4,5 IgG GMCs than non-carriers (Supplementary Table 3).

In the mixture model analyses the 2 estimated distributions overlapped considerably, especially in the O:9 IgA model fit. Iterations of the O:4,5 IgA model did not converge at all and thus we were unable to define a threshold for seroconversion for IgA. The Youden index defined a threshold of 1.15 logAU for O:9 IgG (87% sensitivity and 87% specificity) and 1.45 logAU for O:4,5 IgG (89% sensitivity and 66% specificity). The model fits and ROC analysis are presented in Supplementary [Figure 4-6 and Table 4-5].

Like the GMCs, seroprevalence increased by age across all sites (Figure 3). By site, the highest seroprevalence was among adults in Siaya for both O:9 and O:4,5 IgG. Among 1148 sera tested for both O:9 and O:4,5 IgG, 583 (51%) were seropositive for both antigens while 278 (24%) were negative for both antigens (Supplementary Table 6).

**Figure 3:**
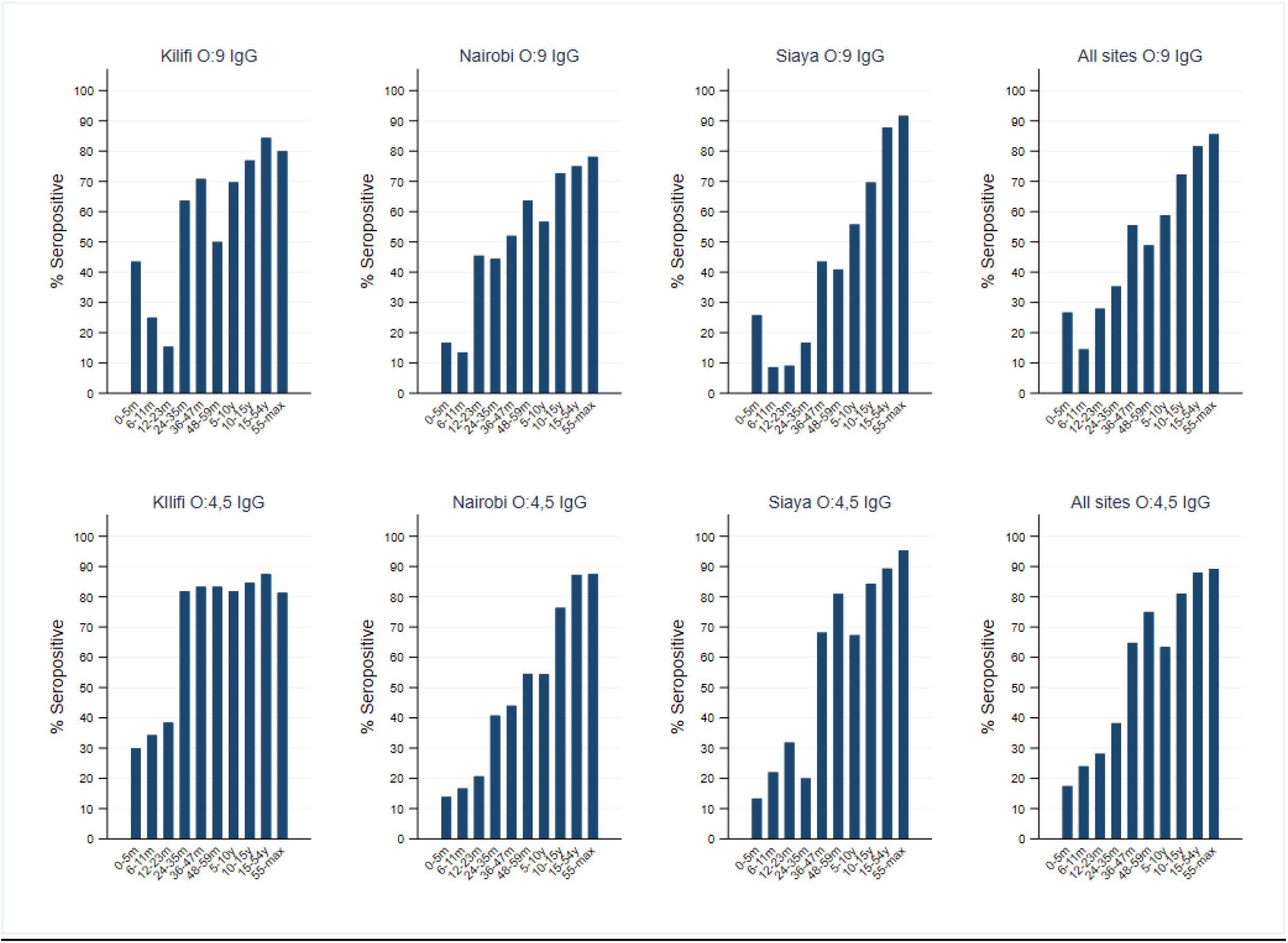
Seroprevalence of O:9 IgG and O:4,5 IgG antibodies by age using Youden’s Index as threshold.

## Discussion

In this randomly sampled population-level sero-survey of NTS, we have observed that both serogroup B (O:4,5) and serogroup D_1_ (O:9) antibodies are present at birth and decay within the first 6 months of life. Endogenous antibodies are acquired in the second half of infancy and increase in concentration and prevalence by age. Maternally derived antibodies to both O:4,5 and O:9 decayed at a rate of 40% per month to very low concentrations in 4-5 months of life, which is similar to observations from Vietnam(14) where maternally-derived anti-NTS antibodies reached their nadir at 20-30 weeks of age. Serum anti-NTS IgA antibodies, remained at low concentrations for the first half of infancy but increased rapidly thereafter, indicating incident infection with bacteria expressing O:4,5 and O:9 antigens.

The lack of IgA responses in early infancy could have several implications. First, it suggests that maternally derived IgG antibodies may have a protective effect against acquisition of infection. Similar observations have been made in cohorts in Vietnam and Malawi where infections and disease, respectively, increased in incidence after maternal IgG antibodies declined; these maternal antibodies exhibited *in vitro* bactericidal activity (14, 29, 30). Lack of seroconversion in early infancy may also be due to reduced exposure during exclusive breastfeeding. However, in Kilifi, the incidence of iNTS disease is highest among neonates (12), especially among those born out of hospital (31). The high incidence of early-onset neonatal iNTS disease could be due to increased mother-to-child transmission of NTS among carrier mothers during vaginal delivery in contaminated environments (31) A maternally administered NTS vaccine could have dual action of reducing mother-to-child transmission of NTS and extending the duration of passive immunity in infants from high-risk areas. In the presence of adequate exposure, a lack of IgA among young infants may also be due to slow maturation of their immune system(32)

We observed an age-related increase in both O:9 and O:4,5 IgG and IgA antibody concentrations and seroprevalence, beginning in second half of infancy in all sites and plateau in late childhood. The rate of increase differed by site reflecting differences in transmission intensities in these endemic regions. For example, the rate of acquisition of O:4,5 IgG was highest in Kilifi; the peak GMC for O:4,5 IgG antibodies was reached by 2 years of age and the seroprevalence was ~70% at 5 years of age, indicating high cumulative incidence of infection with serogroup B *Salmonella* in Kilifi. For O:9 antibodies, the rate of acquisition with age was highest in Nairobi reflecting a high incidence of serogroup D *Salmonella* infections in Nairobi. Interestingly, Kilifi had the highest prevalence of NTS carriage as observed in the parent study (5), including of serogroup B serotypes. The observation in Nairobi is not supported by carriage data, since the single NTS isolated in the carriage study in Nairobi was not serogroup D; neither is it supported by published iNTS disease data which show higher incidence in Siaya than Nairobi(33). It is possible that the O:9 assay was detecting antibodies stimulated by one of the other 165 serotypes within serogroup D, especially *S*. Typhi(34) which is known to cause outbreaks in this urban slum(35).

The relationship between antibody concentrations and faecal carriage of NTS has not been explored previously. We observed higher O:4,5 IgG concentrations among serogroup B carriers than non-carriers, suggesting sustained stimulation and production of antibodies among prevalent carriers.

Whether antibodies protect against acquisition of carriage is not clear, as there is no established threshold of protection against carriage. A longitudinal study would be required to describe the direction of the association between antibody concentrations and faecal carriage of NTS, if causal. In addition, measurement of faecal IgA antibodies may provide a better measure of mucosal immunity to either infection or invasion than IgG and IgA from serum samples(36).

The specificity of the ELISAs used may limit the interpretation of our study. The ELISA was designed to detect the O-antigen, a component of LPS found in multiple gram-negative organisms including NTS. A previous study from Denmark demonstrated cross-reactions with antibodies against other pathogens such as *Escherichia coli, Yersinia enterolitica* and *Campylobacter* sp., in ELISAs targeting the LPS of NTS (37). Additionally, for NTS, multiple serotypes exist within one O-antigen serogroup, and their responses might not be differentiated easily using O-antigen based serology. Our assay-based threshold illustrated this lack of specificity, as it defined seropositivity as any value above the LLOQ. Using this threshold, we observed 100% seroconversion during infancy for both O:4,5 and O:9 IgG across all sites. It is likely that the universal presence of antibodies, using this threshold, simply represents a high background incidence of infection with other gram-negative organisms in the environment including non-pathogenic Salmonellae, and these antibody levels may not be protective against NTS.

The threshold derived from mixture modelling maximized both sensitivity and specificity(28). The utility of the model is contingent on the validity of the underlying assumptions. We made the simplest assumption that the data contained a mixture of 2 populations (seropositive and seronegative) which is the simplest classification possible. Further sub-classifications into recently infected individuals and those with waning antibody could be specified if biologically plausible.

However, as this is one of the first studies to look at population distributions of anti-NTS antibodies, and because our understanding of the responses and waning rates is rudimentary, we selected a two-population model as the most parsimonious option. This approach successfully separated the O:4,5 and O:9 IgG distributions into 2 components each. However, for IgA, the models could not separate any underlying components. The resultant seroprevalence estimates were entirely contingent on the threshold selected. The Youden index (Sensitivity 89% and Specificity 66%) resulted in O:4,5 IgG seroprevalence of 37% in under 5’s while a threshold with 99% specificity and another with 99% sensitivity resulted in seroprevalence estimates that were 36 times lower and 3 times higher, respectively. However, although the thresholds selected here have not been validated externally, they are unlikely to have had a significant impact on the analyses presented here which are focused on the relative comparisons between sites since the same threshold was used at each site.

Without assay standardization, we cannot validate our results through comparisons with other studies. Spatial heterogeneity in our results means that it would be imprudent to generalise the results more widely e.g., to the greater African population. However, we have provided data on key parameters such as prevalence of falciparum malaria, anaemia, and malnutrition, which could be used to adjust the results to compare with other populations. This study provides a perspective on antibody responses after natural infection in Kenya showing that IgG antibody concentrations and prevalence rise sharply throughout childhood from 6 months of age onwards and that the rate of rise varies by setting suggesting different levels of background infection.

## Data Availability

All data produced in the present study are available upon reasonable request to the authors

## Acknowledgements

We thank Dr. Raphael Simon of University of Maryland, USA, for providing the coating antigens for the ELISAs. We thank the field, laboratory and clerical staff of the NTS Project in Kilifi, Nairobi and Siaya, and the study participants.

## Conflict of Interest

No conflict of interest to declare.

## Ethics

The study was approved by the KEMRI Scientific and Ethics Research Unit (SERU No. 3221). This activity was reviewed by CDC and was conducted in a manner consistent with applicable federal law and CDC policy [Project ID: 0900f3eb81e92cdd].

## Funding and Disclaimer

E.M.M is supported through the DELTAS Africa Initiative [DEL-15-003]. The DELTAS Africa Initiative is an independent funding scheme of the African Academy of Sciences (AAS)’s Alliance for Accelerating Excellence in Science in Africa (AESA) and supported by the New Partnership for Africa’s Development Planning and Coordinating Agency (NEPAD Agency) with funding from the Wellcome Trust and the UK government. S.F was supported by a Sir Henry Dale Fellowship jointly funded by the Wellcome Trust and the Royal Society (Grant number 208812/Z/17/Z). C.A.M. was supported by an MRC Senior Clinical Fellowship (MR/R005974/1) J.A.G.S is supported by a Wellcome Trust Senior Fellowship (214320). The funders had no role in study design, data collection and analysis, decision to publish, or preparation of the manuscript. The findings and conclusions in this report are those of the authors and do not necessarily represent the official position of the U.S. Centers for Disease Control and Prevention. This article was published with the approval of the KEMRI Director.

## Notes

### Competing Interest Statement

The authors have declared no competing interest.

### Author Declarations

The KEMRI Scientific and Ethics Research Unit gave ethical approval for this work (SERU No. 3221)

## REFERENCES

1. Collaborators GN-TSID. The global burden of non-typhoidal salmonella invasive disease: a systematic analysis for the Global Burden of Disease Study 2017. Lancet Infectious Diseases. 2019;19:1312–24.

2. Hagedoorn NN, Murthy S, Birkhold M, Marchello CS, Crump JA. Prevalence and distribution of non-typhoidal Salmonella enterica serogroups and serovars isolated from normally sterile sites: A global systematic review. Epidemiology and Infection. 2024;152:e4.

3. Feasey NA, Dougan G, Kingsley RA, Heyderman RS, Gordon MA. Invasive non-typhoidal salmonella disease: an emerging and neglected tropical disease in Africa. Lancet (London, England). 2012;379(9835):2489–99.

4. Gal-Mor O. Persistent Infection and Long-Term Carriage of Typhoidal and Nontyphoidal Salmonellae. Clinical microbiology reviews. 2019;32(1).

5. Muthumbi EM, Mwanzu A, Mbae C, Bigogo G, Karani A, Mwarumba S, et al. The epidemiology of fecal carriage of nontyphoidal Salmonella among healthy children and adults in three sites in Kenya. PLOS Neglected Tropical Diseases. 2023;17(10):e0011716.

6. Im J, Nichols C, Bjerregaard-Andersen M, Sow AG, Løfberg S, Tall A, et al. Prevalence of Salmonella Excretion in Stool: A Community Survey in 2 Sites, Guinea-Bissau and Senegal. Clinical infectious diseases : an official publication of the Infectious Diseases Society of America. 2016;62 Suppl 1(Suppl 1):S50–5.

7. Haselbeck AH, Im J, Prifti K, Marks F, Holm M, Zellweger RM. Serology as a Tool to Assess Infectious Disease Landscapes and Guide Public Health Policy. Pathogens [Internet]. 2022; 11(7).

8. Arnold B, Scobie H, Priest J, Lammie P. Integrated Serologic Surveillance of Population Immunity and Disease Transmission. Emerging Infectious Disease journal. 2018;24(7):1188.

9. Nyiro JU, Kombe IK, Sande CJ, Kipkoech J, Kiyuka PK, Onyango CO, et al. Defining the vaccination window for respiratory syncytial virus (RSV) using age-seroprevalence data for children in Kilifi, Kenya. PLOS ONE. 2017;12(5):e0177803.

10. Stephens DS. Protecting the herd: the remarkable effectiveness of the bacterial meningitis polysaccharide-protein conjugate vaccines in altering transmission dynamics. Trans Am Clin Climatol Assoc. 2011(0065-7778 (Print)).

11. Smith PG. Concepts of herd protection and immunity. Procedia in Vaccinology. 2010;2(2):134–9.

12. Muthumbi E, Morpeth SC, Ooko M, Mwanzu A, Mwarumba S, Mturi N, et al. Invasive Salmonellosis in Kilifi, Kenya. Clinical Infectious Diseases. 2015;61(Suppl_4):S290–S301.

13. Voysey M, Kelly DF, Fanshawe TR, Sadarangani M, O’Brien KL, Perera R, et al. The Influence of Maternally Derived Antibody and Infant Age at Vaccination on Infant Vaccine Responses : An Individual Participant Meta-analysis. JAMA Pediatrics. 2017;171(7):637–46.

14. de Alwis R, Tu LTP, Quynh NLT, Thompson CN, Anders KL, Van Thuy NT, et al. The Role of Maternally Acquired Antibody in Providing Protective Immunity Against Nontyphoidal Salmonella in Urban Vietnamese Infants: A Birth Cohort Study. The Journal of Infectious Diseases. 2019;219(2):295–304.

15. Kuhn KG, Falkenhorst G, Ceper TH, Dalby T, Ethelberg S, Mølbak K, et al. Detecting non-typhoid Salmonella in humans by ELISAs: a literature review. Journal of Medical Microbiology. 2012;61(1):1–7.

16. Weber GE, White MT, Babakhanyan A, Sumba PO, Vulule J, Ely D, et al. Sero-catalytic and Antibody Acquisition Models to Estimate Differing Malaria Transmission Intensities in Western Kenya. Scientific Reports. 2017;7(1):16821.

17. Pothin E, Ferguson NM, Drakeley CJ, Ghani AC. Estimating malaria transmission intensity from Plasmodium falciparum serological data using antibody density models. Malaria Journal. 2016;15(1):79.

18. Chan Y, Fornace K, Wu L, Arnold BF, Priest JW, Martin DL, et al. Determining seropositivity—A review of approaches to define population seroprevalence when using multiplex bead assays to assess burden of tropical diseases. PLOS Neglected Tropical Diseases. 2021;15(6):e0009457.

19. Sepúlveda N, Stresman G, White MT, Drakeley CJ. Current Mathematical Models for Analyzing Anti-Malarial Antibody Data with an Eye to Malaria Elimination and Eradication. Journal of Immunology Research. 2015;2015(1):738030.

20. Biggs JR, Sy AK, Sherratt K, Brady OJ, Kucharski AJ, Funk S, et al. Estimating the annual dengue force of infection from the age of reporting primary infections across urban centres in endemic countries. BMC Medicine. 2021;19(1):217.

21. Bottomley C, Otiende M, Uyoga S, Gallagher K, Kagucia EW, Etyang AO, et al. Quantifying previous SARS-CoV-2 infection through mixture modelling of antibody levels. Nature Communications. 2021;12(1):6196.

22. Idris ZM, Chan CW, Kongere J, Hall T, Logedi J, Gitaka J, et al. Naturally acquired antibody response to Plasmodium falciparum describes heterogeneity in transmission on islands in Lake Victoria. Scientific Reports. 2017;7(1):9123.

23. Elias SC, Muthumbi E, Mwanzu A, Wanjiku P, Mutiso A, Simon R, et al. Complementary measurement of nontyphoidal Salmonella-specific IgG and IgA antibodies in oral fluid and serum. Heliyon. 2023;9(1):e12071.

24. Baliban SM, Yang M, Ramachandran G, Curtis B, Shridhar S, Laufer RS, et al. Development of a glycoconjugate vaccine to prevent invasive Salmonella Typhimurium infections in sub-Saharan Africa. PLOS Neglected Tropical Diseases. 2017;11(4):e0005493.

25. Stockdale L, Nalwoga A, Nash S, Elias S, Asiki G, Kusemererwa S, et al. Cross-sectional study of IgG antibody levels to invasive nontyphoidal Salmonella LPS O-antigen with age in Uganda [version 1; peer review: 2 approved, 2 approved with reservations]. Gates Open Research. 2019;3(1501).

26. Cox MJ, Azevedo RS, Cane PA, Massad E, Medley GF. Seroepidemiological study of respiratory syncytial virus in São Paulo State, Brazil. Journal of Medical Virology. 1998;55(3):234–9.

27. Deb P. FMM: Stata module to estimate finite mixture models.. Boston College Department of Economics;. 2012.

28. Youden WJ. Index for rating diagnostic tests. (0008-543X (Print)).

29. MacLennan CA, Gondwe EN, Msefula CL, Kingsley RA, Thomson NR, White SA, et al. The neglected role of antibody in protection against bacteremia caused by nontyphoidal strains of Salmonella in African children. The Journal of Clinical Investigation. 2008;118(4):1553–62.

30. Nyirenda TS, Gilchrist JJ, Feasey NA, Glennie SJ, Bar-Zeev N, Gordon MA, et al. Sequential Acquisition of T Cells and Antibodies to Nontyphoidal Salmonella in Malawian Children. The Journal of Infectious Diseases. 2014;210(1):56–64.

31. Talbert AWA, Mwaniki M, Mwarumba S, Newton CRJC, Berkley JA. Invasive Bacterial Infections in Neonates and Young Infants Born Outside Hospital Admitted to a Rural Hospital in Kenya. The Pediatric Infectious Disease Journal. 2010;29(10).

32. Wood N, Siegrist C-A. Neonatal immunization: where do we stand? Current Opinion in Infectious Diseases. 2011;24(3).

33. Verani JR, Toroitich S, Auko J, Kiplang’at S, Cosmas L, Audi A, et al. Burden of Invasive Nontyphoidal Salmonella Disease in a Rural and Urban Site in Kenya, 2009–2014. Clinical Infectious Diseases. 2015;61(Suppl_4):S302–S9.

34. Grimont P, Weill F. Antigenic Formulae of the Salmonella serovars. 2008.

35. Kariuki S, Revathi G, Kiiru J, Mengo Doris M, Mwituria J, Muyodi J, et al. Typhoid in Kenya Is Associated with a Dominant Multidrug-Resistant Salmonellaenterica Serovar Typhi Haplotype That Is Also Widespread in Southeast Asia. Journal of Clinical Microbiology. 2010;48(6):2171–6.

36. Ndungo E, Pasetti MF. Functional antibodies as immunological endpoints to evaluate protective immunity against Shigella. Human Vaccines & Immunotherapeutics. 2020;16(1):197–205.

37. Strid Mette A, Dalby T, Mølbak K, Krogfelt Karen A. Kinetics of the Human Antibody Response against Salmonella enterica Serovars Enteritidis and Typhimurium Determined by Lipopolysaccharide Enzyme-Linked Immunosorbent Assay. Clinical and Vaccine Immunology. 2007;14(6):741–7.

