## Supplementary Material for "The age-specific concentrations and seroprevalence of antibodies against *Salmonella* Enteritidis (O:9) and *Salmonella* Typhimurium (O:4,5) across three sites in Kenya"

**List of Tables and Figures.**

Table 1. Number of samples tested for each antigen-antibody combination.

Table 2: Participant characteristics among the subset of participants with complete results on all four isotype-serotype combinations tested (n=1,148).

Table 3A: Geometric mean concentrations of O:9 IgG and IgA antibodies among those with serogroup D carriage and those without serogroup specific carriage.

Table 3B: Geometric mean concentrations of O:4,5 IgG and IgA antibodies among those with serogroup B carriage and those without serogroup specific carriage.

Table 4: Predicted class means and associated class proportions from mixture modelling.

Table 5: Sensitivity and Specificity of different cut-offs.

Table 6: Association between seroprevalence of O:9 IgG and seroprevalence of O:4,5 IgG

Figure 1A: Scatterplots of antibody concentration by age in years with LOWESS curves fitted: O:9 IgG and IgA

Figure 1B: Scatterplots of antibody concentration by age in years with LOWESS curves fitted: O:4,5 IgG and IgA

Figure 2A: Scatterplots of antibody concentration by age in years (<10y) with LOWESS curves fitted: O:9 IgG and IgA

Figure 2B: Scatterplots of antibody concentration by age in years (<10y) with LOWESS curves fitted: O:4,5 IgG and IgA

Figure 3: Scatter plot with LOWESS showing crude rate of decay in maternal antibodies by age at each site.

Figure 4: Histograms showing the observed vs predicted densities from a 2-component mixture model

Figure 5: Receiver Operating Characteristic (ROC) Curve of O:9 and O:4,5 IgG concentrations.

Figure 6: Seroprevalence of O:9 and O:4,5 IgG by age and site, Lower limit of quantification cut-off.

Table 1. Number of samples tested for each antigen-antibody combination.

| | O:9 IgG | O:4,5 IgG | O:9 IgA | O:4,5 IgA |
|---|---|---|---|---|
| Vials retrieved | 1254 | 1254 | 1254 | 1254 |
| Insufficient Volume | 1 | 2 | 53 | 55 |
| Failed CV criterion (>20) | 1 | 12 | 7 | 5 |
| Plate Failed* | - | 22 | - | - |
| | 1252 | 1216 | 1194 | 1194 |

*($R^2$<0.994 or Controls out of range)

Table 2: Participant characteristics among the subset of participants with complete results on all four isotype-serotype combinations tested (n=1,148).

| | Kilifi | | Nairobi | | Siaya | | Total | |
|---|---|---|---|---|---|---|---|---|
| | n | % | n | % | n | % | n | % |
| N | 302 | | 397 | | 449 | | 1,148 | |
| Age | | | | | | | | |
| 0-11m | 50 | 17 | 54 | 14 | 73 | 16 | 177 | 15 |
| 12-59m | 58 | 19 | 78 | 20 | 96 | 21 | 232 | 20 |
| 5-14y | 59 | 20 | 137 | 35 | 102 | 23 | 298 | 26 |
| 15-54y | 62 | 21 | 97 | 24 | 75 | 17 | 234 | 20 |
| >55y | 73 | 24 | 31 | 8 | 103 | 23 | 207 | 18 |
| Sex, male | 123 | 41 | 198 | 50 | 197 | 44 | 518 | 45 |
| MUAC <11.5cm in under 5y† | 1 | 0.9 | 22 | 17 | 5 | 3 | 28 | 7 |
| Number tested for Hb concentration* | 291 | 96 | 347 | 87 | 213 | 47 | 851 | 74 |
| Anaemia (Hb<10g/dl)* | 57 | 20 | 17 | 5 | 19 | 9 | 93 | 11 |
| Number tested for *P. falciparum* malaria* | 295 | 98 | 357 | 90 | 441 | 98 | 1,093 | 95 |
| *P. falciparum* positive by HRP2* | 21 | 7 | 9 | 3 | 122 | 28 | 152 | 14 |

MUAC – Mid Upper Arm Circumference

† Only measured for children under 5years of age

*Number tested for Hb or HRP-2 as a subset of the total participants

The median age (and interquartile range) for the participants were 11y (3-54) in Kilifi, 9y (3-23) in Nairobi, 10y (3-48) in Siaya and 9.7y (2.8-34) for all sites combined

Table 3A: Geometric mean concentrations of O:9 IgG and IgA antibodies among those with serogroup D carriage and those without serogroup specific carriage.

| | | Carriers of serogroup D | | Carriers of other serogroups | | No carriage | | GMC Ratio | |
|---|---|---|---|---|---|---|---|---|---|
| Antibody | Age | n | GMC (95% CI) | n | GMC(95% CI) | n | GMC(95% CI) | Serogroup D vs other serogroup | Serogroup D vs No carriage |
| O:9 IgG | <5 years | 3 | 27 (0, 4324) | 5 | 8 (1-52) | 466 | 6 (5-8) | 3.3 (0-747) | 4.2 (0.5-37.1) |
| | 5+ years | 3 | 55(2.4, 1277) | 25 | 53 (25-116) | 749 | 41 (36-46) | 1.0 (0.1-10.3) | 1.4 (0.2-8.1) |
| O:9 IgA | <5 years | 2 | 27 (0-1500) | 5 | 8 (3-19) | 427 | 7 (6-8) | 3.6 (0.3-37.8) | 3.8 (0.7-22.5) |
| | 5+ years | 3 | 38 (8-190) | 25 | 34 (23-51) | 731 | 37 (34-40) | 1.1 (0.3-3.8) | 1.0 (0.3-3.9) |

Table 3B: Geometric mean concentrations of O:4,5 IgG and IgA antibodies among those with serogroup B carriage and those without serogroup specific carriage.

| | | Carriers of serogroup B | | Carriers of other serogroups | | No carriage | | GMC Ratio | |
|---|---|---|---|---|---|---|---|---|---|
| Antibody | Age | n | GMC | n | GMC | n | GMC | Serogroup B vs other serogroup | Serogroup B vs No carriage |
| O:4,5 IgG | <5 years | - | - | 7 | 14 (2-88) | 447 | 11 (9-14) | - | - |
| | 5+ years | 10 | 177 (74-424) | 18 | 88 (47-163) | 733 | 72 (66-79) | 2.0 (0.7-5.5) | 2.4 (1.1-5.4) |
| O:4,5 IgA | <5 years | - | - | 7 | 6 (2-16) | 426 | 5 (4-6) | - | - |
| | 5+ years | 10 | 59 (16-212) | 18 | 27 (12-61) | 732 | 40 (36-44) | 2.2 (0.5-8.7) | 1.5 (0.7-3.3) |

Figure 1A: Scatterplots of antibody concentration by age in years with LOWESS curves fitted: O:9 IgG and IgA

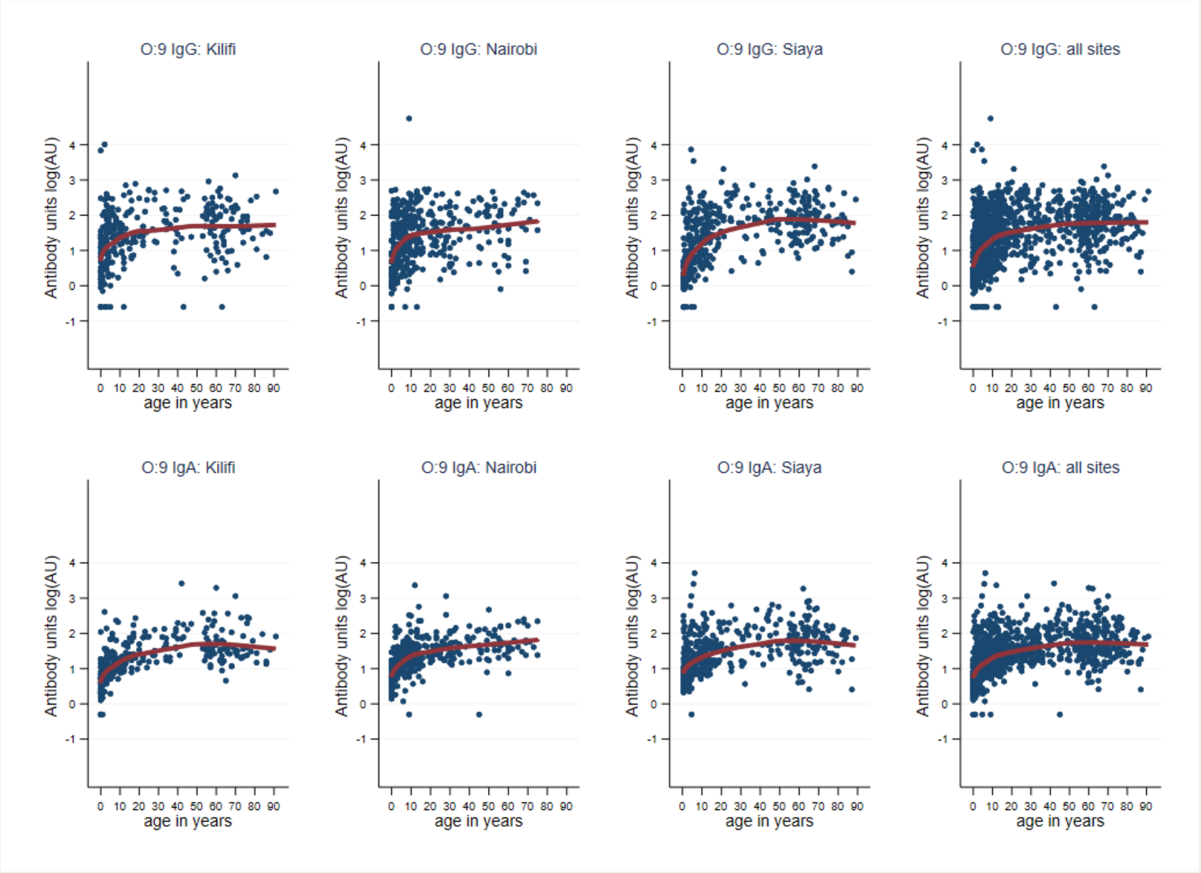

Figure 1B: Scatterplots of antibody concentration by age in years with LOWESS curves fitted: O:4,5 IgG and IgA

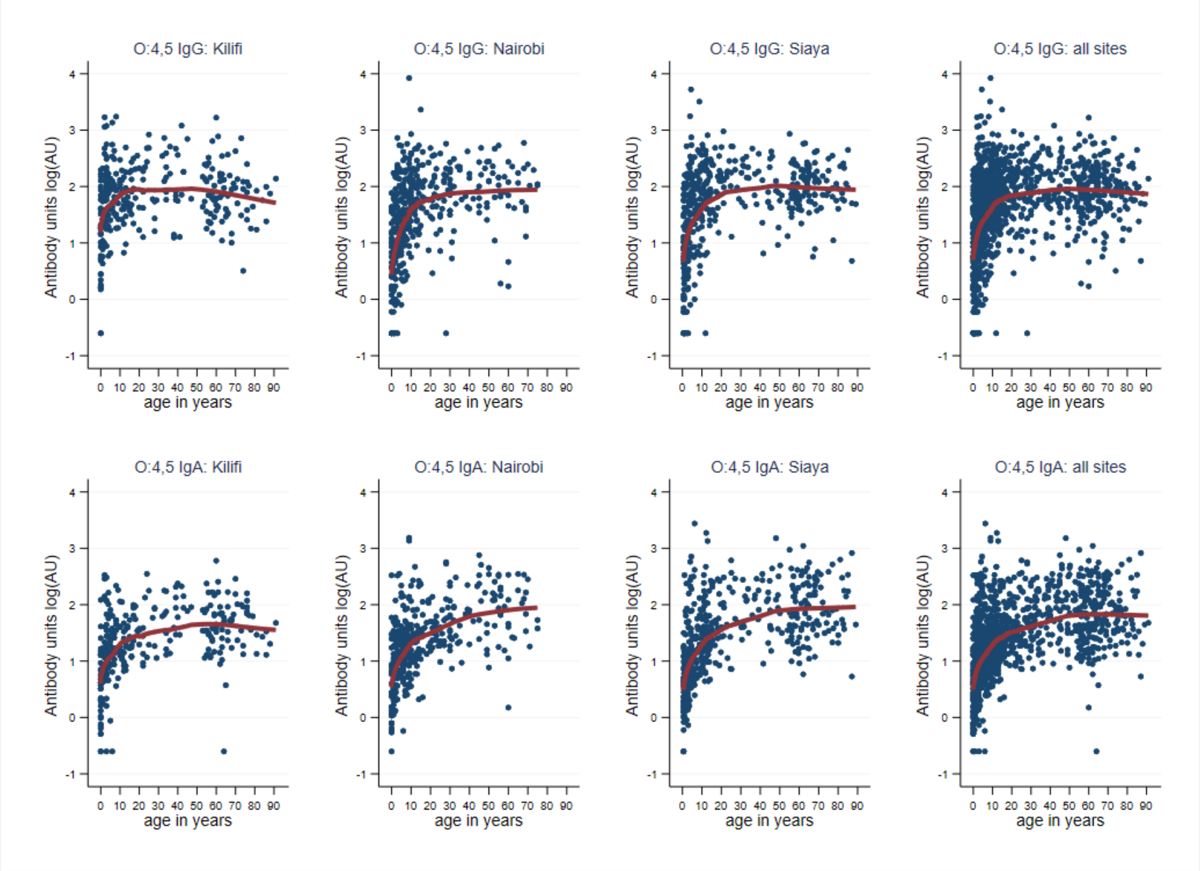

Figure 2A: Scatterplots of antibody concentration by age in years (<10y) with LOWESS curves fitted: O:9 IgG and IgA

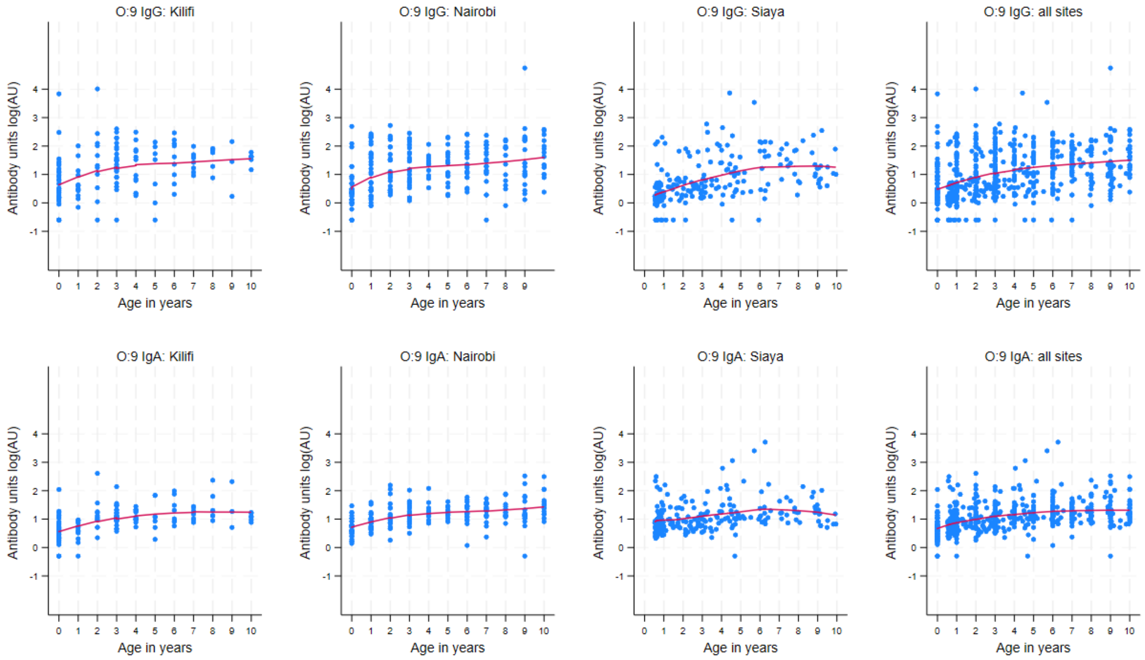

Figure 2B: Scatterplots of antibody concentration by age in years (<10y) with LOWESS curves fitted: O:4,5 IgG and IgA

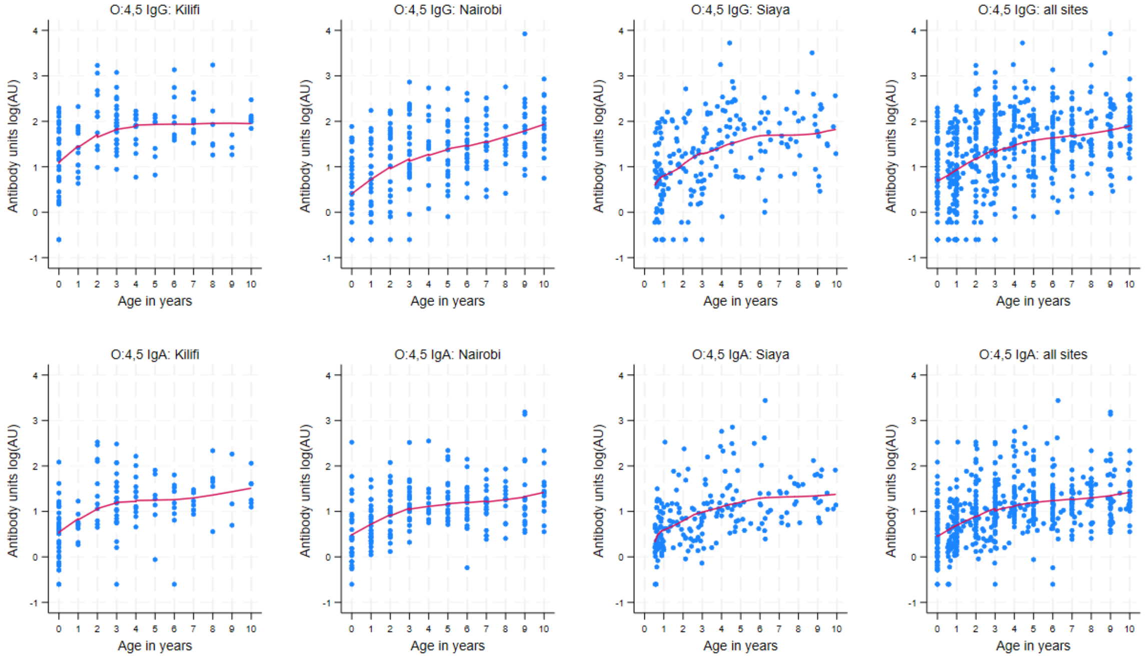

Figure 3: Scatter plot with LOWESS showing crude rate of decay in maternal antibodies by age at each site.

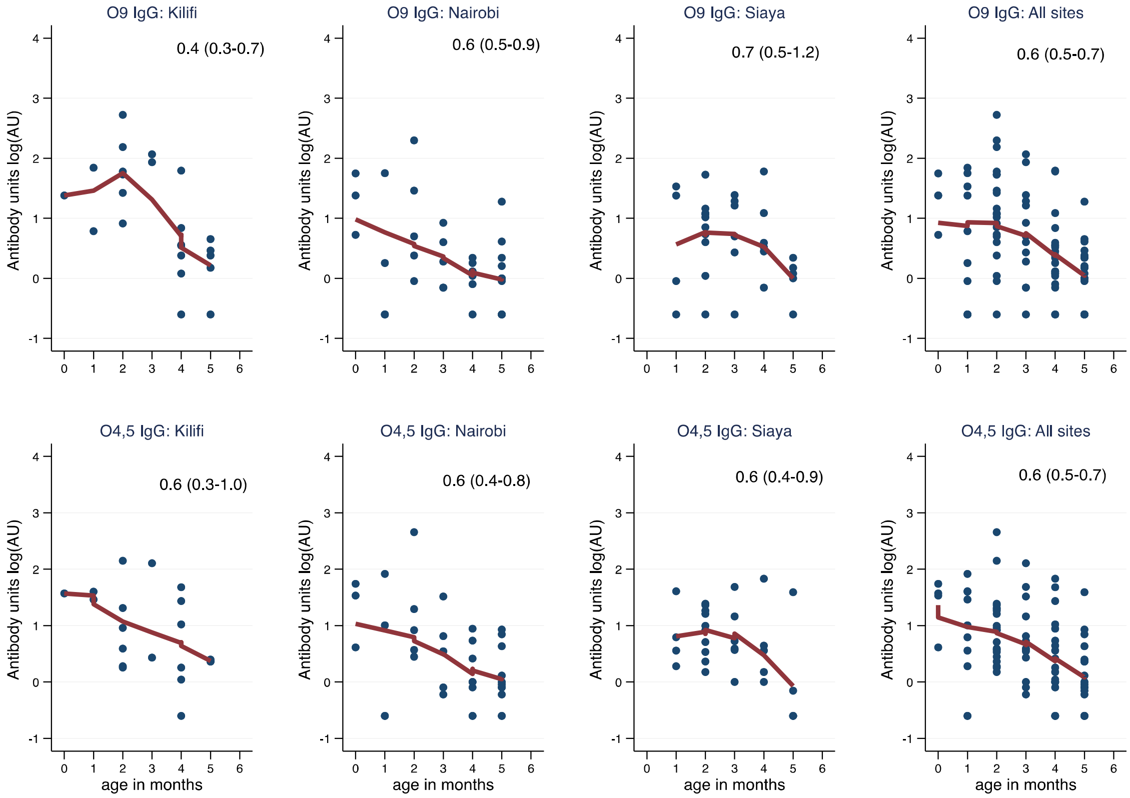


The numbers represent the crude rate of decay (and 95% CI) estimated for each site and serotype by linear regression.

Figure 4: Histograms showing the observed vs predicted densities from a 2-component mixture model

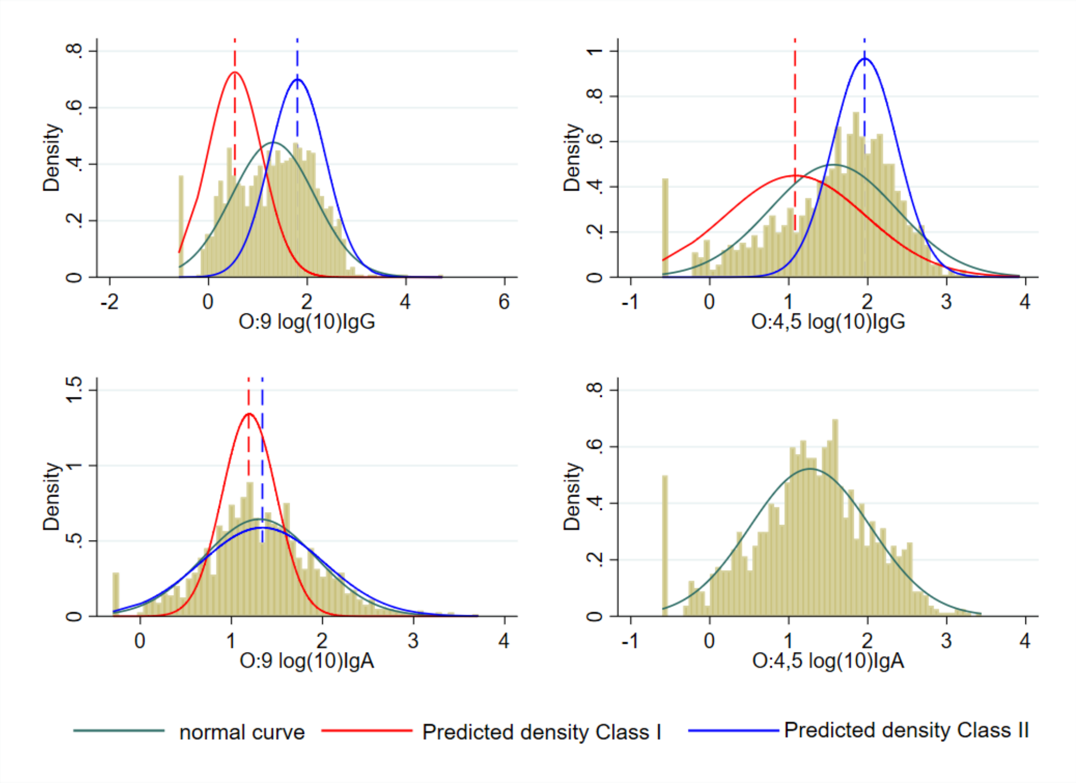

Table 4: Predicted class means and associated class proportions from mixture modelling.

| | O:9 IgG | | O:4,5 IgG | | O:9 IgA | |
|---|---|---|---|---|---|---|
| | Class 1 | Class 2 | Class 1 | Class 2 | Class 1 | Class 2 |
| Predicted Mean (Log AU) | 0.53 | 1.8 | 1.08 | 1.96 | 1.19 | 1.34 |
| (SD) | (0.5) | (0.6) | (0.9) | (0.4) | (0.3) | (0.7) |
| Predicted Proportion | 0.38 | 0.61 | 0.45 | 0.55 | 0.22 | 0.78 |
| (95% CI) | (0.27-0.51) | (0.48-0.72) | (0.36-0.54) | (0.45-0.64) | (0.08-0.44) | (0.56-0.92) |

Figure 5: Receiver Operating Characteristic (ROC) Curve of O:9 and O:4,5 IgG concentrations.

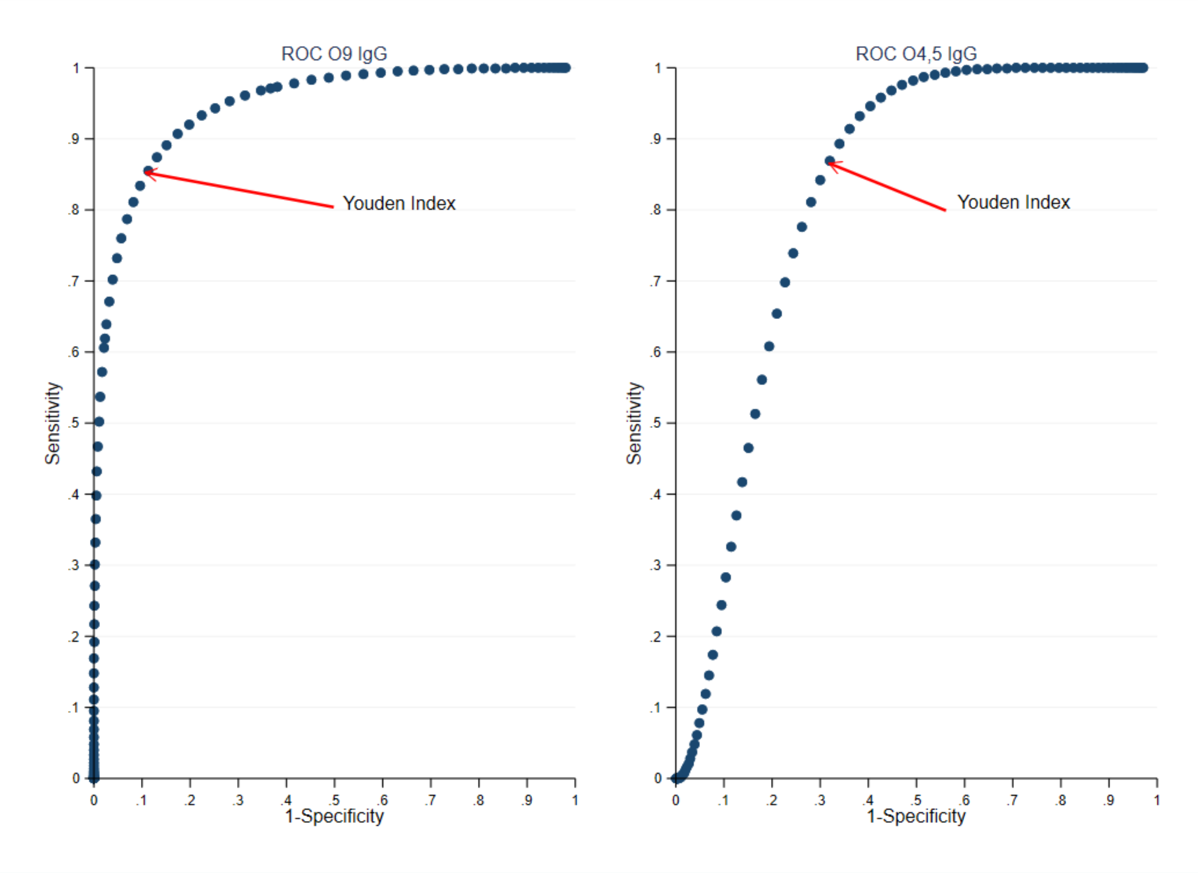

Table 5: Sensitivity and Specificity of different cut-offs.

| | O:9 IgG | | | O:4,5 IgG | | |
|---|---|---|---|---|---|---|
| | Mean of Class 1 + 2SD | Youden Index | LLOQ | Mean of Class 1 + 2SD | Youden Index | LLOQ |
| *Thresholds* | | | | | | |
| Cut off LogAU | 1.63 | 1.15 | -0.3 | 2.88 | 1.45 | -0.3 |
| Cut off AU | 42.7 | 14.1 | 0.5 | 758.6 | 28.2 | 0.5 |
| *Test statistics* | | | | | | |
| Sensitivity | 62% | 87% | 99% | 1% | 89% | 99% |
| Specificity | 97% | 87% | 48% | 98% | 66% | 7% |
| *Seroprevalence* | | | | | | |
| 0-11 m | 21/218 (10%) | 43/218 (20%) | 191/218 (87%) | 0/204 (0%) | 44/204 (22%) | 176/204 (86%) |
| 12-59 m | 65/256 (25%) | 105/256 (41%) | 250/256 (98%) | 5/250 (2%) | 123/250 (49%) | 240/250 (96%) |
| 5-14 y | 125/312 (40%) | 202/312 (65%) | 307/312 (98%) | 6/307 (2%) | 218/307 (71%) | 306/307 (100%) |
| 15-54 y | 146/250 (58%) | 204/250 (82%) | 249/250 (100%) | 4/241 (2%) | 212/241 (88%) | 240/241 (100%) |
| 55+ y | 133/215 (62%) | 184/215 (86%) | 214/215 (100%) | 3/213 (1%) | 190/213 (89%) | 213/213 (100%) |

Figure 6: Seroprevalence of O:9 and O:4,5 IgG by age and site, Lower limit of quantification cut-off.

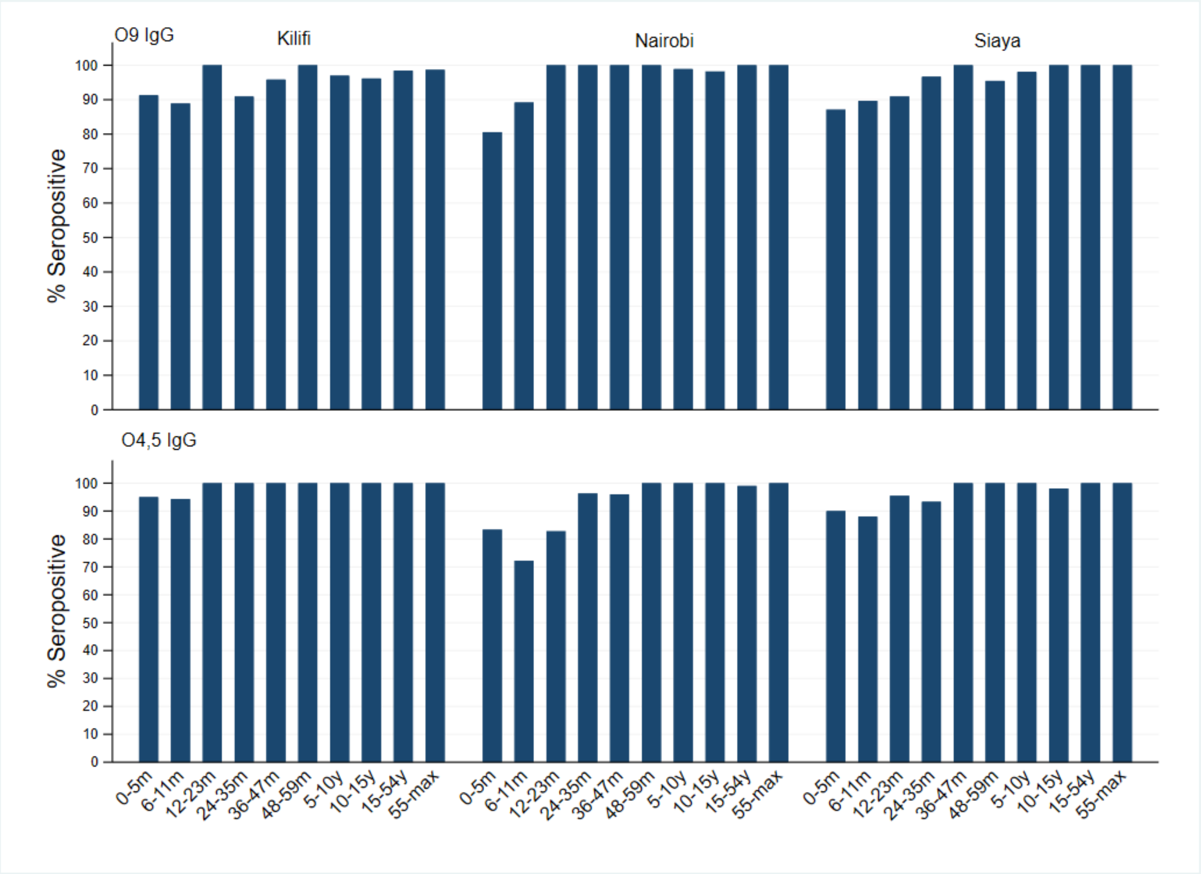

Table 6: Association between seroprevalence of O:9 IgG and seroprevalence of O:4,5 IgG

| | Kilifi | Nairobi | Siaya | All |
|---|---|---|---|---|
| N (number of paired samples) | 302 | 397 | 449 | 1148 |
| Seropositive for both antibodies | 170 (56%) | 175 (44%) | 238 (53%) | 583 (51%) |
| Seronegative for both antibodies | 48 (16%) | 104 (26%) | 126 (28%) | 278 (24%) |
| O:4,5 seropositive, O:9 seronegative | 52 (17%) | 63 (16%) | 63 (14%) | 178 (16%) |
| O:9 seropositive, O:4,5 seronegative | 32 (11%) | 55 (14%) | 22 (5%) | 109 (9%) |
| McNemar $c^2$ | 4.76 | 0.54 | 19.78 | 16.6 |
| p-value | 0.03 | 0.461 | <0.001 | <0.001 |

seroprevalence differed significantly in Siaya and Kilifi but not in Nairobi, meaning the risk factors for 0:9 and 0:4,5 infections are not shared in Nairobi, while in Kilifi and Siaya the two serogroups have shared risk factors.
